# Tool for estimating the probability of having COVID-19 with one or more negative RT-PCR results

**DOI:** 10.1101/2021.01.16.21249939

**Authors:** Alejandro Jara, Eduardo A. Undurraga, Rafael Araos

**Affiliations:** Department of Statistics, Pontificia Universidad Católica de Chile, Santiago, RM, Chile; Millenium Nucleus Center for the Discovery of Structures in Complex Data (MIDaS) Santiago, RM, Chile; Escuela de Gobierno, Pontificia Universidad Católica de Chile, Santiago, RM, Chile; Millennium Initiative for Collaborative Research in Bacterial Resistance (MICROB-R), Santiago, RM, Chile; Instituto de Ciencias e Innovación en Medicina, Universidad del Desarrollo, Santiago, RM, Chile

**Author notes:** **Address for correspondence**: Alejandro Jara, Department of Statistics, Pontificia Universidad Católica de Chile, Av. Vicuña Mackenna 4860, Macul CP 7820436, Santiago, Región Metropolitana, Chile.

**Keywords:** COVID-19, RT-PCR test, Bayesian, online tool

## Abstract

Early case detection and isolation of infected individuals are critical to controlling COVID-19. RT-PCR is considered the diagnosis gold standard, but false-negatives occur. Based on previous work, we built a user-friendly online tool to estimate the probability of having COVID-19 with negative RT-PCR results and thus avoid preventable SARS-CoV-2 transmission.

---

The COVID-19 pandemic has imposed a catastrophic toll worldwide, with about 66 million reported cases and 1.5 million reported deaths as of December 4, 2020 [1]. Despite the historic approval of a COVID-19 vaccine tested in a large clinical trial in the United Kingdom [2], epidemic control still critically depends on non-pharmaceutical interventions, such as social distancing, and early detection and effective isolation of infected individuals [3].

Compared to other viruses, SARS-CoV-2, the virus that causes COVID-19 disease, has two characteristics that make early case detection and isolation of infected individuals particularly critical for epidemic control. First, the virus is an efficient spreader, with an average of about 2.5 secondary infections caused by a single infected individual in a susceptible population [4]. Second, the highest risk of transmission occurs very early in the disease, before or within the first days of symptom onset [5]. These characteristics hinder transmission control because early case detection and effective isolation are challenging. Thus far, COVID-19 surveillance has been primarily based on reverse transcriptase-polymerase chain reaction (RT-PCR) assays, considered the most reliable diagnostic test for COVID-19 [6]. However, RT-PCR positivity varies among infected patients depending on, for example, the timing of sample collection in relation to symptom onset or the sampling technique used (e.g., nasopharyngeal swabs, sputum) [7].

Accurate testing results are critical to prevent onward SARS-CoV-2 transmission in the community and hospital settings. As the second wave of infection continues to grow in Europe and the United States, straining health system capacity even further, critical workers with known exposures to the virus will continue to use RT-PCR tests to discard SARS-CoV-2 infection. False-negative RT-PCR results are particularly problematic among healthcare and other essential workers (e.g., firefighters, police), which may inadvertently become “superspreaders” [8]. As social distancing measures are relaxed, RT-PCR false-positives also affect community control measures if workers with suspected infection are cleared to return to work. If negative RT-PCR results are treated as evidence of no-infection, there is a non-negligible risk of preventable transmission of the virus.

We addressed this problem by designing a readily available, easy-to-use online tool to estimate the probability that an individual is infected with SARS-CoV-2 conditional on having one or more negative RT-PCR test results (https://midas-uc.shinyapps.io/Calculadora-COVID19/). Our tool, based mainly on the work by Kucirka et al. [9], requires users to first choose between one, two, or three consecutive negative RT-PCR tests. Second, to provide (i) the estimated probability of being positive before taking the RT-PCR diagnostic test (range: 0 to 1), and (ii) the number of days elapsed between the first symptoms and the first, second, or third RT-PCR test (range: -4 to 16 days). The tool is available in English and Spanish.

We build on Kucirka et al.’s [9] work and adjusted a hierarchical logistic model to estimate the rate of false negatives for different moments in time from the onset of symptoms. We implemented the model using JAGS and the RJAGS library in R [10]. We generated a Markov chain of 420,000 samples; the first 20,000 samples were discarded, and the rest were re-sampled to generate a sub-chain of size 20,000. In contrast to Kucirka et al. [9], we estimated a non-study-specific marginal rate of false-negative RT-PCR tests. Our online tool assumes a specificity of one of the RT-PCR test for detecting SARS-CoV-2 and independence of the results of different tests when considering more than one test (specific details are shown in the Technical Appendix).

To show the tool’s utility, we input data from a healthcare worker in a hospital setting in Chile who was not working with COVID-19 patients. The worker was inadvertently exposed to SARS-CoV-2 by an asymptomatic patient who later developed symptoms and had confirmed COVID-19. The worker had a negative nasopharyngeal swab RT-PCR result three days following exposure. She reported symptoms the day after the first negative RT-PCR and had a second RT-PCR test that showed positive results (Technical Appendix). Because she was exposed for a relatively long period, we used a 50% pre-test probability of infection (Figure).

**Figure.**
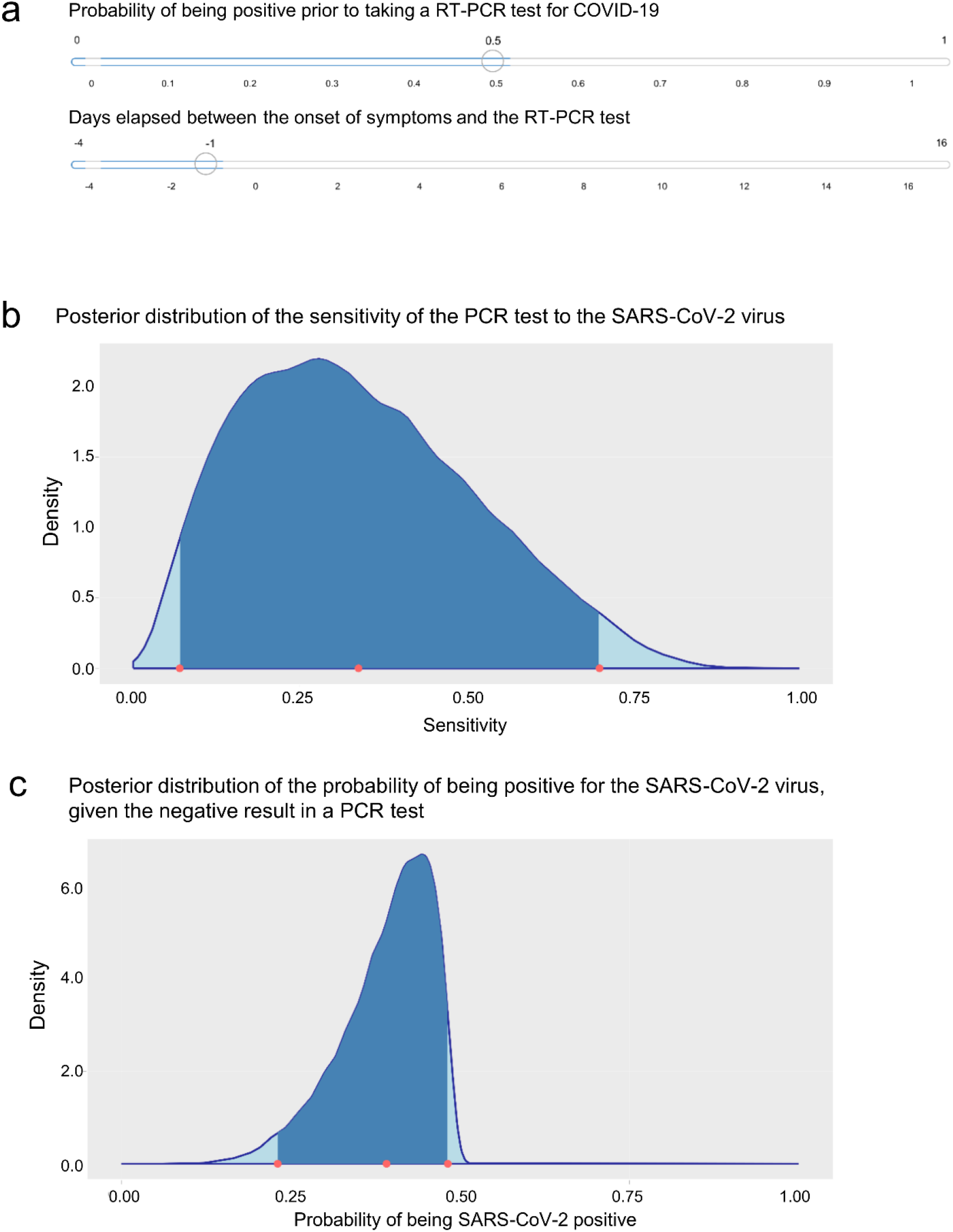
Illustrative results from the tool to estimate the probability of having COVID-19 with one negative RT-PCR tool. (a) The worker reported symptoms the day after the first negative RT-PCR; because she was exposed for a relatively long period of time we used a 50% pre-test probability of infection (b) shows the posterior distribution of the sensitivity of the PCR test to the SARS-CoV-2 virus, and (c) shows the posterior distribution of the probability of being positive for the SARS-CoV-2 virus, given the negative result in a PCR test

Our online tool enables users to estimate the probability of having COVID-19 with one or more negative RT-PCR results. The tool is available in English and Spanish. We hope this publicly available online tool will help decision-makers to avoid preventable transmission of SARS-CoV-2.

## Data Availability

All data are publicly available

https://midas-uc.shinyapps.io/Calculadora-COVID19/

## Acknowledgments

This work was supported by the ANID Millennium Science Initiative grants MIDAS NCN17_059 and MICROB-R NCN17_081.

## Technical appendix

### Tool for estimating the probability of having COVID-19 with one or more negative RT-PCR results

Our online tool is based on Kucirka et al. [9]. In Kucirka et al.’s article, the data of samples from the upper respiratory tract of n = 1330 patients reported in seven studies on the performance of the PCR test for the detection of the SARS-CoV-2 virus at different moments of the disease development [11-17] were analyzed.

Kucirka et al. [9] considered only nasal samples in their analysis and adjusted a hierarchical logistic model to estimate the rate of false negatives for different moments in time from the onset of symptoms. Let *Y*_*ij*_ be the number of positive patients, out of a total of *n*_*ij*_, in the study *i, i* = 1, …, 7, at day *j* since exposure to SARS-COV-2. Kucirka et al. [9] considered the hierarchical model given by

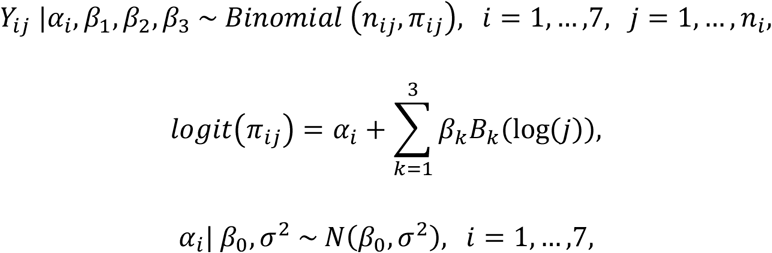

where *B*_*k*_ is the function that evaluates the orthogonal polynomial of degree *k, β*_1_, …, *β*_3_ are regression coefficients, *α*_*i*_ is the random effect of study *i, i* = 1, …, 7, *N*(*μ, τ*) denotes the normal distribution with mean *μ* and variance τ. Kucirka et al. [9] implemented a Bayesian version of the model using the STAN library of the statistical program R [10]. They estimated the rate of false negatives on the day *j*, based on the expression

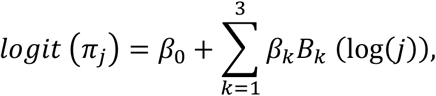

which generates a conditional (study-specific) estimate of the false-negative rate.

We partially reproduced the analysis carried out by Kucirka et al. [9]. Specifically, we considered the same data and hierarchical model, which was implemented using the following prior distributions:

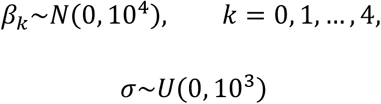

We implemented the model using the JAGS program [18] and the RJAGS library [19] https://cran.r-project.org/web/packages/rjags/index.html of the R statistical program [10]. We generated a Markov chain of 420,000 samples; the first 20,000 samples were discarded, and the rest were re-sampled to generate a sub-chain of size 20,000.

Unlike Kucirka et al.’s [9] work, we estimated the false-negative rate using the expression

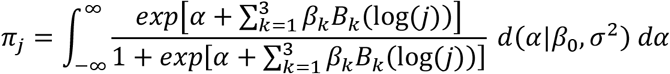

where *d*(· | *μ, σ*^2^) denotes the density of the normal distribution with mean *μ* and variance *σ*^2^, which generates an estimate of the marginal rate of false negatives (not study-specific). Figure S1 shows the posterior median and the 95% credibility interval limits for the false-negative rate.

**Figure S1.**
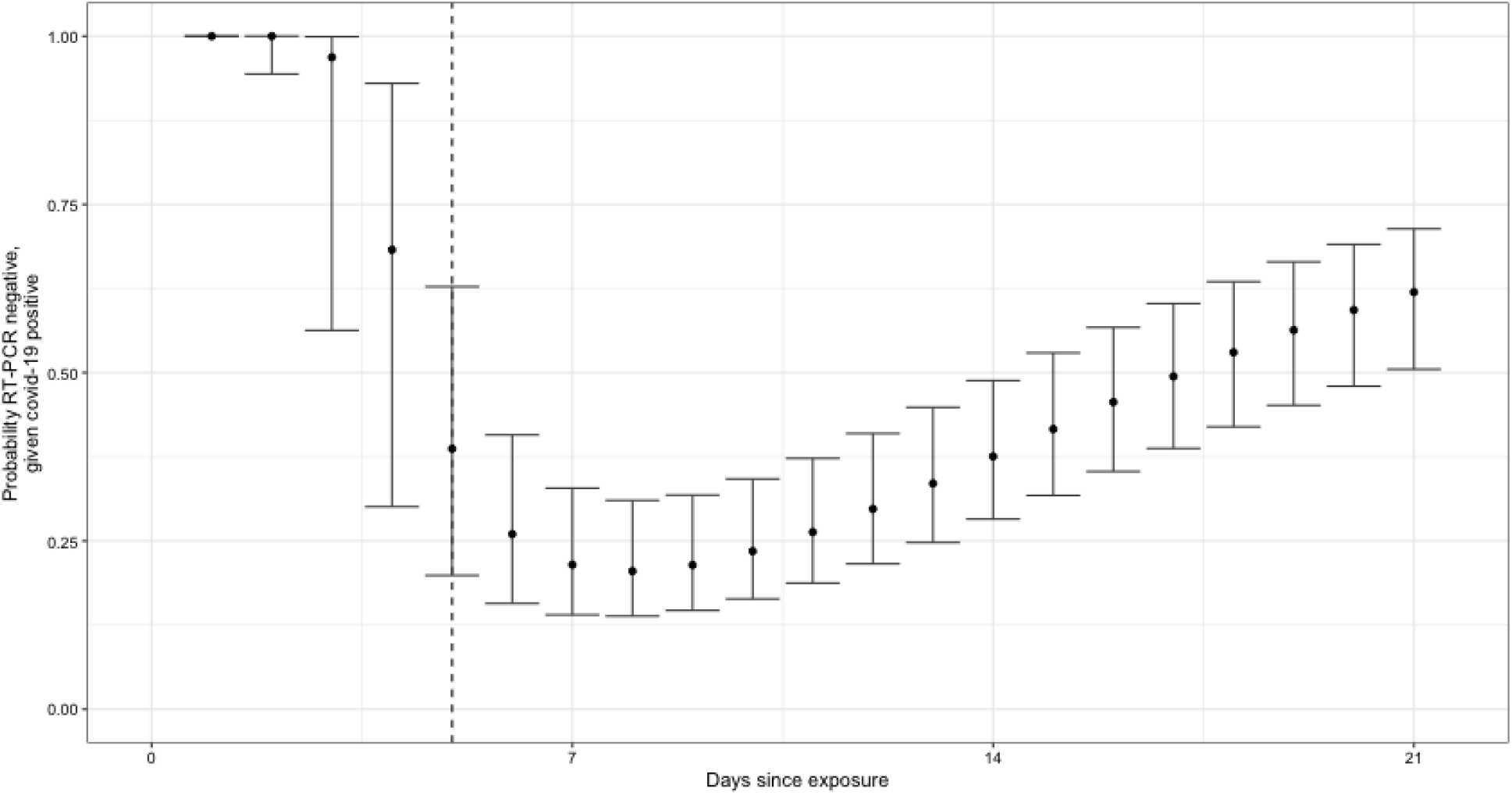
Posterior median and 95% credibility intervals for the false-negative rate of PCR tests by day since exposure to SARS-CoV-2

Our online tool assumes a specificity of one of the PCR tests for detecting SARS-CoV-2 virus and independence of the results of different tests when considering more than one test. The tool is publicly available in English and Spanish at https://midas-uc.shinyapps.io/Covid19-calculator/ and https://midas-uc.shinyapps.io/Calculadora-COVID19/, respectively.

